# Unlocking the Power of EHRs: Harnessing Unstructured Data for Machine Learning-based Outcome Predictions

**DOI:** 10.1101/2023.02.13.23285873

**Authors:** Mohammad Noaeen, Somayeh Amini, Shveta Bhasker, Zohreh Ghezelsefli, Aisha Ahmed, Omid Jafarinezhad, Zahra Shakeri Hossein Abad

## Abstract

The integration of Electronic Health Records (EHRs) with Machine Learning (ML) models has become imperative in examining patient outcomes due to the vast amounts of clinical data they provide. However, critical information regarding social and behavioral factors that affect health, such as social isolation, stress, and mental health complexities, is often recorded in unstructured clinical notes, hindering its accessibility. This has resulted in an over-reliance on clinical data in current EHR-based research, potentially leading to disparities in health outcomes. This study aims to evaluate the impact of incorporating patient-specific context from unstructured EHR data on the accuracy and stability of ML algorithms for predicting mortality, using the MIMIC III database. Results from the study confirmed the significance of incorporating patient-specific information into prediction models, leading to a notable improvement in the discriminatory power and robustness of the ML algorithms. Furthermore, the findings underline the importance of considering non-clinical factors related to a patient’s daily life, in addition to clinical factors, when making predictions about patient outcomes. These results have significant ramifications for improving ML in clinical decision support and patient outcome predictions.

## I. Introduction

Clinical factors have a limited influence on a patient’s overall health outcome, accounting for only 10-20%. The rest is largely shaped by social, environmental, and behavioral factors [1]–[3]. Despite the vast amount of health data comprising 30% of the world’s information, healthcare systems continue to struggle with a lack of information and resulting disparities in access and equity. The Social and Behavioral Determinants of Health (SBDoH), which are largely overlooked in outcome research, play a crucial role in enhancing the inclusiveness and quality of healthcare systems. For example, wealthier Canadians are generally healthier and live longer than poorer Canadians [4]. The obesity rate is 1.5 times higher for Canadian women in lower-income households than higher-income families (20% versus 13%), putting them at higher risks of diabetes and coronary artery disease [5]. Canadians with the lowest incomes report less heavy drinking but are over two times as likely to be hospitalized for conditions entirely caused by alcohol [6]. This highlights the need to consider the influence of additional factors such as inadequate nutrition, lack of social support, or elevated stress levels in our assessments.

The widespread adoption of Electronic Health Records (EHRs) has generated large volumes of clinical data. This is an enabling resource for developing machine learning (ML) models to study patient outcomes (e.g. [7]–[11]). However, information about social determinants of health is usually recorded in unstructured clinical notes, which hampers access to this information. Thus, most current ML-based outcome research using EHRs focuses heavily on clinical factors and consequentially may lead to health inequalities. A recent study highlights the impact of uncertainty on the performance of EHR-based mortality prediction models among critically ill patients and underscores the importance of a comprehensive understanding of individual patient contexts to evaluate the robustness and generalizability of ML-based models [12].

The integration of social and behavioral information into EHRs is an area of growing interest in the healthcare industry [13]–[20]. Pantell et al. [18] investigated the correlation between social and behavioral elements and the earlier development of hypertension and diabetes among a clinical sample. The results of their cohort study involving 18,133 adults without baseline hypertension and 35,788 adults without baseline diabetes shows that cumulative social and behavioral risk factors, such as low education level and infrequent exercise, are significantly linked to earlier onset of both hypertension and diabetes. In a similar study, Yu et al. [19] compared the information obtained from Natural Language Processing (NLP)-extracted SBDoH with structured EHRs in a cohort of 864 patients and found that more in-depth information about factors like smoking, education, and employment was only present in the clinical narratives. This confirms the necessity of utilizing both clinical narratives and structured EHRs to gain a comprehensive understanding of a patient’s health conditions. Feller et al. [20] used NLP to extract SBDoH that impact the prediction of HIV diagnosis. They found that the inclusion of these factors significantly improved the predictive performance of automated HIV risk assessment by identifying terms in clinical text indicative of high-risk behavior.

In the context of mental health, studies have shown that individuals with mental health conditions, such as depression and anxiety, are at a higher risk of death from physical health conditions, including cardiovascular disease and cancer [21], [22]. Furthermore, there is a significant association between pre-existing depression and mortality [23]. To further explore this and to evaluate the potential usefulness of incorporating unstructured text data from EHRs for improved personalized patient information extraction, we utilized patient notes from the Medical Information Mart for Intensive Care III (MIMIC III) database [24] to extract information related to mental health conditions, including stress, anxiety, depression, and post-traumatic stress disorder (PTSD), and to examine the predictive capability of these factors on patient mortality outcomes. Our study reveals the pivotal role that patient-specific information plays in enhancing the accuracy of our machine learning models in predicting the mortality outcome. Our findings not only confirm the importance of incorporating mental health information into mortality predictions, but also highlight the benefits of utilizing a comprehensive dataset that incorporates both laboratory data and SBDoH in training and evaluating the ML models.

## II. Methods

### A. Data Collection and Preparation

#### Dataset

The present study utilized data sourced from the Medical Information Mart for Intensive Care III (MIMIC III) database, which is a collaborative effort between Philips Healthcare and the Laboratory for Computational Physiology at the Massachusetts Institute of Technology (MIT) [24]. The database comprises information from 53,423 hospital admissions of adult patients over the age of 16 who were admitted to critical care units between 2001 and 2012, as well as data from 7,870 neonatal admissions between 2001 and 2008. The MIMIC III database encompasses 38,597 unique adult patients and 49,785 hospital admissions, encompassing various data types, including vital signs, medications, and laboratory measurements. To reduce the impact of prior ICU admissions on patients with multiple stays in the ICU, only the last stay was considered in the analysis. However, to gain a comprehensive understanding of the patient’s mental health conditions recorded in the unstructured notes, we merged and incorporated all of the patient’s clinical notes into the labeling process.

#### Data Labeling

A sample of 1,058 patient records was carefully selected and assigned a binary mental health label (0 or 1) by experienced research assistants who are specialists in the fields of psychology, clinical epidemiology, and nursing. Each record was meticulously reviewed by the research assistants, who examined the historical clinical notes and based their labeling on the reports provided by the clinical care team. This label was then incorporated into the 25-variable MIMIC III dataset, which encompasses patient demographics and first 24-hour ICU physiological and lab variables. As mental health can affect individuals of all ages, no age range was established as a criterion for exclusion. To ensure a balanced representation of both death and non-death cases, equal distribution of these cases was maintained in the sample dataset.

In addition to manually labeling the clinical notes to extract social and behavioral factors, we also leveraged the ChatGPT model as a data annotation tool. We utilized OpenAI’s GPT-3 Playground and its programmatic API to identify the presence of mental health issues in the unstructured notes. The model annotated 900 records, and the results were compared with the manual labeling process.

#### Data Preprocessing

To address the issue of missing data, we employed the use of missingness indicators in our models. This approach ensured the preservation of the original distribution of features and reduced the loss of data records. Additionally, we standardized the numerical features by converting them into *z*-scores. This was accomplished by subtracting the mean and scaling each feature to have unit variance. Moreover, our correlation analysis revealed that Chloride and Sodium as well as Hematocrit and Hemoglobin have a strong correlation with each other, with a correlation coefficient greater than 95%. To minimize the risk of over-fitting, we chose to exclude Sodium and Hemoglobin as they had a higher number of missing data compared to Chloride and Hematocrit.

### B. Prediction Models Development

To mitigate over-fitting and ensure accurate results with regards to our limited dataset, we evaluated a wide range of standard machine learning classifiers with varying architectures, including Logistic Regression (LR), kernel-based Support Vector Machines (SVM), decision-tree-based Random Forest (RF), XGBoost, ExtraTrees, and sample-based K-Nearest Neighbors (KNN). To gauge the influence of mental health context information extracted from clinical notes on the performance of these models, we implemented and compared the outcomes of two different settings: (1) predictions based solely on clinical data and (2) predictions augmented with mental health information extracted from unstructured text data. To ensure the robustness and generalizability of our results, we applied 5-fold cross validation in our evaluation process. The performance of each classifier was measured based on several key metrics, including precision, recall, F1 scores, and the Area Under the Curve (AUC). To prioritize the classifier’s ability to detect positive cases (i.e., death outcome), we also employed average precision (AP), which places a higher emphasis on the precision-recall trade-off.

## III. Results and Discussion

### Mortality Prediction

The performance of our predictive models in predicting mortality is presented in Table I. The models were trained using raw clinical data and data augmented with additional context on the patient’s mental health conditions. With the exception of the kernel-based SVM classifier, the performance of other classifiers either improved or remained consistent when trained on the augmented dataset compared to the raw dataset. The ExtraTrees classifier applied to the augmented dataset outperformed the other models in our study, showing a 4% improvement in precision, recall, F1, and average precision (AP) metrics compared to its performance on the raw data (i.e., Precision: 81%, Recall: 81%, F1: 81%). The highest AP score of 75% and AUC score of 89% (Figure 1) were achieved using the ExtraTrees classifier on the augmented dataset. The lower performance of SVM when trained on the augmented dataset may be due to its sensitivity to the dimensionality of the data. As the number of dimensions in the dataset increases, SVM becomes less effective in finding the optimal boundary that separates the classes.

**TABLE I:**
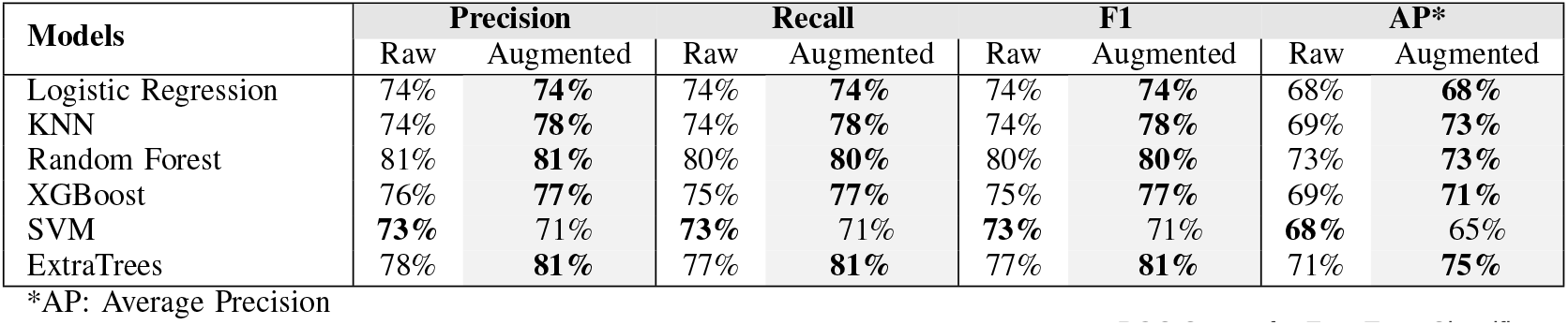
Comparison of the raw and augmented datasets for different predictive models. The Augmented column displays the performance of the models that were trained and tested on the combination of clinical data and unstructured data.

**Fig. 1:**
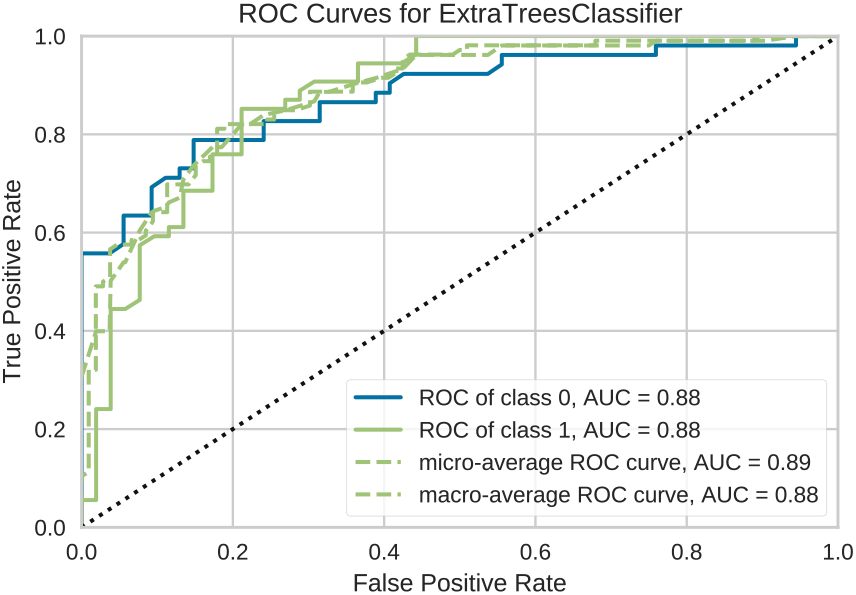
Two-class classification AUC for the dataset augmented with mental health information (ExtraTrees).

### Mental Health Contribution

Figure 2 (a,b) shows the contribution of the top 20 features to the performance of our best-performing model (ExtraTrees). We utilized SHapley Additive exPlanations (SHAP) analysis [25] to assess the contribution of each feature to individual predictions. Our results showed that the binary mental health feature had a significant effect on the predictions made by the augmented model, ranking 13th among the top 20 features despite the limited sample size, as depicted in Figure 2a. The summary plot in Figure 2b uses red to indicate a positive contribution of the feature to the prediction, which increases the prediction from its baseline value, and blue to represent a negative contribution, which decreases the prediction from its baseline value. For instance, a high positive contribution is seen for higher values of age, while this is the reverse for the platelet feature. The binary nature of the augmented mental health information is evident in the final prediction, which is impacted by either value of this feature, highlighting the importance of incorporating patient-specific context into prediction models.

**Fig. 2:**
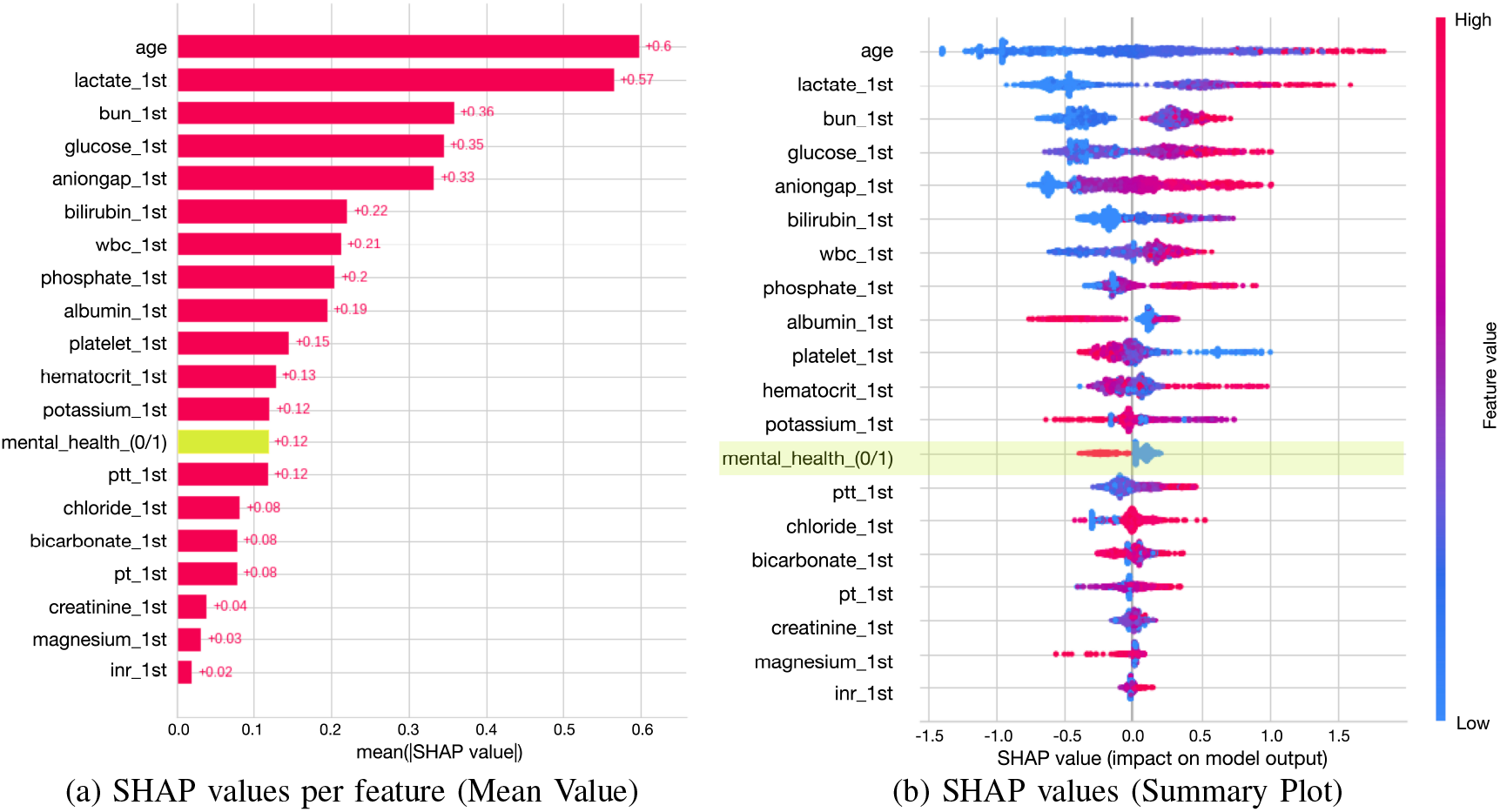
The contribution of each feature to the prediction of mortality using the augmented dataset for the best-performing model (ExtraTrees). The values on the x-axis show (a) the mean absolute and (b) the impact values.

### OpenAI and Data Labeling

In order to evaluate the reliability and practicality of utilizing the OpenAI API for detecting SBDoH from unstructured text, we compared the labels generated by the AI to those provided by domain experts. Our results revealed an agreement of 77% between the two sources of labels. Nevertheless, only 5 out of 111 positive labels (indicating the presence of mental health) were accurately detected by OpenAI, and the majority of the agreements with domain experts were in the negative category. This implies that the AI module, despite its large language model, may require further adjustments to achieve a higher level of accuracy for this task. Additionally, the intricacy of detecting mental health and the varied nature of the concepts related to this health condition could account for the high level of discrepancy between the outputs of the AI and domain experts in labeling positive cases.

### Study Limitations

This study has several limitations that may affect the validity of our findings. Firstly, the small sample size of the dataset used in our research may impact the generalizability of our results. To mitigate this, we have taken measures to ensure high quality label extraction and a balanced random selection of the dataset to reduce the impact of small sample size and imbalanced data. Secondly, while our study focuses solely on mental health-related SBDoH, these factors often interact with one another and should be considered in aggregate. To address this, a team of four domain experts conducted a thorough review of each patient’s clinical notes to account for interactions among SBDoH that may impact mental health. Despite these limitations, our results contribute to the field of outcome research and highlight the importance of incorporating patient-specific context in EHR-based outcome studies using machine learning.

## IV. Conclusion

In this study, we sought to examine the impact of incorporating mental health information from SBDoH into unstructured notes in EHRs on the accuracy of mortality prediction using machine learning. We used this information to generate and validate prediction models that can be utilized to identify high-risk patients, taking into account both clinical and interrelated social factors. Our findings reveal that integrating mental health data from clinical notes enhances the performance of mortality prediction models compared to models relying solely on clinical data. Furthermore, the augmented information extracted from the clinical notes was found to be a key contributing factor in predicting mortality outcomes for our best-performing machine learning model. This underscores the significance of incorporating SBDoH and patient-specific context into EHR-based outcome analysis.

## Data Availability

We used MIMIC III dataset, which requires its own credentials to access.

## ACKNOWLEDGMENT

This project is funded by the Data Science Institute– Computational & Quantitative Social Sciences (CQSS), University of Toronto.

